# Efficient and accurate simulation of infectious diseases on adaptive networks

**DOI:** 10.1101/2024.12.02.24318307

**Authors:** Nils Gubela, Max von Kleist

## Abstract

Mathematical modelling of infectious disease spreading on temporal networks has recently gained popularity in complex systems science to understand the intricate interplay between social dynamics and epidemic processes. While analytic solutions for these systems can usually not be obtained, numerical studies through exact stochastic simulation has remained infeasible for large, realistic systems.

Here, we introduce a rejection-based stochastic sampling algorithm with high acceptance probability (‘high-acceptance sampling’; HAS), tailored to simulate disease spreading on adaptive networks. We proof that HAS is exact and can be multiple orders faster than Gillespie’s algorithm. While its computational efficacy is dependent on model parameterization, we show that HAS is applicable regardless on whether contact dynamics are faster, on the same time-scale, or slower than the concurrent disease spreading dynamics. The algorithm is particularly suitable for processes where the spreading- and contact processes are co-dependent (adaptive networks), or when assumptions regarding time-scale separation become violated as the process unfolds. To highlight potential applications, we study the impact of diagnosis- and incidence-driven behavioural changes on virtual Mpox- and COVID-like epidemic and examine the impact of adaptive behaviour on the spreading processes.

**Author Summary:** Infectious disease spreading is often affected by the dynamics of human-human contacts. These contact dynamics may change over time, and in direct response to infection kinetics, through e.g. self-isolation, risk-aversion, or any *adaptive* behaviour, which can generate complex dynamics as seen in recent outbreaks with e.g. COVID-19, as well as Mpox clade IIb (2022). Agent-based models (ABMs) are often derived and numerically simulated to study the complex interplay between epidemic- and contact dynamics and to derive insights for disease control. However, numerical simulation of these models denotes a computational bottleneck and limits the applicability of large ABMs. We introduce a novel numerical method called ‘high-acceptance sampling’ (HAS), which allows for the exact simulation of outbreaks with *adaptive* contact behaviour. We proof that HAS is exact, show that it is faster, and that runtime grows with at least an order of magnitude less than state-of-the art exact simulation methods. This enables simulation of outbreaks on large populations, as well as parameter estimation for large systems. We apply HAS to study an Mpox- and COVID-like pandemic and the impact of adaptive behaviour on different time-evolving contact networks.

## 1 Introduction

Mathematical modelling and simulation has a long history in epidemiology [1, 2, 3]. In recent outbreaks like SARS-CoV-2 or Mpox, mathematical modelling has contributed decisively to guiding and understanding public health interventions [4, 5], disease prevalence [6], as well as pathogen evolution [5]. However, many approaches have also been developed because of convenient mathematical properties that are not satisfied in a real life setting [7]. For example, mean-field models are easy to analyse but do not capture relevant spreading paths and may provide incorrect predictions [8]. In addition, mean-field models are unable to adequately capture adaptive social behavior [9].

In recent years, agent-based models have become increasingly popular in epidemiology and the social sciences [10, 11, 12, 13, 14, 15]. Agents are discrete, autonomous entities that behave according to simple rules. Agent-based models are easy to understand, but can capture complex dynamics, e.g. through socio-behavioural rules. In our case, we model the spread of a pathogen between heterogeneous agents. The agents are nodes in a time-dependent adaptive contact network, which can be understood as a social network or a sexual contact network, depending on the pathogen [16, 17, 18].

The spread of the pathogen between two connected agents and the dynamics of the contact network are described by stochastic processes. The spreading process depends on the state of the network, since transmission is only possible between connected agents. Importantly, the evolution of the network also depends on the state of the infection process [19]. For example, diagnosed agents may isolate themselves and meet fewer people until they recover. Similarly, a critical number of infected agents may induce risk-averse behaviour in susceptible agents. A convenient assumption for the analysis of dynamical processes in evolving networks is the separation of time-scales between the contact network process and the spreading process. If the network process is much faster than the spreading process, the connectivity patterns effectively disappear and classical mean-field or degree-based ordinary differential equation models may be appropriate approximations to the original system [20]. If the spreading process is much faster than the network process, only a static snapshot of the network is observed by the spreading process. In this case, either static network modelling [21, 22] or modelling of the contact network dynamics may be sufficient to approximate epidemic dynamics [23, 24, 25]. Both assumptions allow the use of a wide range of analytical and numerical methods. However, in most real-world applications the network and the spreading process evolve on comparable time-scales [26, 27, 28, 29, 30, 31].

The gold standard for exact simulation of agent-based systems is the Gillespie or stochastic sampling algorithm (SSA) [32, 33, 34, 35]. It is a continuous-time Markov algorithm in which a single simulation step follows a Poisson process and the duration of that step is therefore sampled from a real-valued waiting time distribution. An update is either an epidemic event or a change in the contact network, which makes the SSA algorithm unsuitable for large networks.

There are already adaptations of SSA to networks: For example, if the dynamics of the contact network are independent of the spreading process, a realisation of the temporal network can be precomputed [23]. The (now fixed) temporal network can then be used for exact simulations of the spreading process [36, 37, 38], similar to the ‘integral method’ used in systems biology [39, 40]. However, in realistic settings, contact topology and spreading dynamics co-evolve, which is referred to as ‘adaptive networks’ [16, 41, 42, 43, 44]. For adaptive networks in particular, none of the previously introduced approaches can be used because each simulated trajectory has its own realisation of the network, which must be simulated in parallel. Some implementations therefore arbitrarily discretise time and perform parallel updates of the contact network and contagion spreading [45, 46], leading to numerical errors that depend on the chosen time discretisation [47]. For large time steps, computationally efficient simulations can be achieved, but the introduced numerical errors make the simulation results unreliable [48]. On the other hand, small time steps offer no computational advantage over the SSA [49]. Finally, it is completely unclear a priori which time step is appropriate as the dynamics and relevant time scales of the process may change over the course of the simulation.

We use a network model where the presence of each edge over time is governed by a continuous-time Markov chain. This allows us to configure the expected degree of each node per time step, as well as properties like assortativity, degree correlations, and clustering, resulting in realistic network structures. When all rates are homogeneous, low-dimensional approximations for ‘edge classes’ can be derived [50, 51], which are numerically fast to solve and analytically tractable.

We formulate an exact rejection-based algorithm to simulate spreading dynamics on adaptive networks as a generalisation of the method of St-Onge et al. [52]. The core idea is to focus on epidemic events and leap over most individual contact updates. Because we design the method as a high acceptance sampling (HAS) algorithm, it is much faster than SSA in all real-world applications, yet it generates trajectories from the same statistical distributions as SSA. HAS is applicable to adaptive network models involving all combinations of network and spreading time scales and requires no assumptions in regards to mixing of nodes (agents). Furthermore, the network process can have several different time scales. We compare the performance of HAS with SSA and highlight the influence of adaptivity on various epidemic spreading models. Proofs of the exactness of HAS can be found in the supplementary materials.

## 2 Methods

We model the spread of infection in a population by representing the population as a temporal network and allowing the infection to propagate through it. This process involves network events, contact-dependent events (such as infections), and spontaneous transition events.

### 2.1 The network model

In our model, we examine a system with *N* agents that dynamically form and dissolve contacts with each other over time, leading to a heterogeneous network. Each agent is represented as a node in a graph, and an edge is drawn between two nodes when the corresponding agents are in contact. The time-dependent adjacency matrix ***A***(*t*) has an entry *a*_*ij*_(*t*) = 1 if agent *i* is in contact with agent *j* at time *t*, and *a*_*ij*_(*t*) = 0 otherwise. The status of each edge *a*_*ij*_ is governed by a two-state continuous-time Markov process, where edges are created at rate 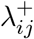 and removed at rate 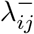. These rates are expressed in inverse time units.

To scale the time of the system, we fix 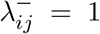, ensuring that each connection persists for an average duration of one time unit. Each agent possesses its own pair of contact parameters, 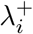 and 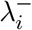, which are used to determine the edge rates 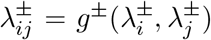 [53]. For example, 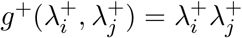 creates a neutral network, where nodes connect randomly, while 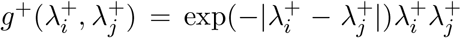 generates an assortative network, where nodes with similar degree tend to connect. In summary, we obtain the reactions 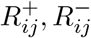

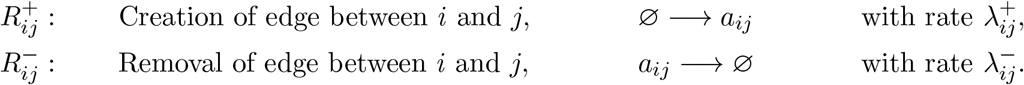

The rates can be calculated from a distribution ***μ*** that represents the expected number of contacts over the simulation time frame [0, *T*]. The probability of creating a link between agent *i* and agent *j* during the simulation is given by 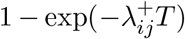, hence the rates can be determined by solving

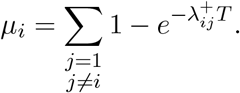

This equation ensures that, throughout the simulation, the expected number of contacts for each agent aligns with the specified distribution. The expected number of contacts per agent should remain constant, *μ* _*i*_ = *O*(1), rather than scaling with the size of the system. This implies that the creation rates scale inversely with the number of agents, 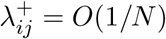 (since exp(−1*/N*) *>* 1 − 1*/N*).

### 2.2 Contact-driven agent-based contagion models

In the simplest form, the spreading is modeled by dividing each agent into one of two compartments, either susceptible *S* or infected *I*. Upon contact, a susceptible agent *i* may become infected by an infected agent at a rate 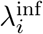. This reaction is dependent on the relationship of two agents and hence we call it contact dependent event. While this reaction is fundamental to all spreading models, it can be expanded by incorporating additional spontaneous transitions. These transitions do not rely on contacts and affect only a single agent at a time. Adaptivity is introduced by considering the diagnosis of infected agents, thereby adding a diagnosed compartment *D*. The infected agent *i* transitions to the D compartment with rate 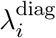. Once diagnosed, agent *i* loses all existing contacts and forms new contacts at a reduced rate, 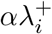, where *α* ∈ (0, 1). This system will be referred to as SID. We also consider models in which the infectious period eventually ends and infected or diagnosed agents become recovered. Agents in the recovered compartment *R* are not eligible for transmitting infection or becoming infectious. Recovery occurs with rate 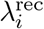 and we refer to this model as SIDR. Lastly, we consider the case in which recovered agents could become susceptible again (SIDRS). The spontaneous transition from *R* to *S* occurs at rate 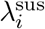. In summary, we consider the following reactions

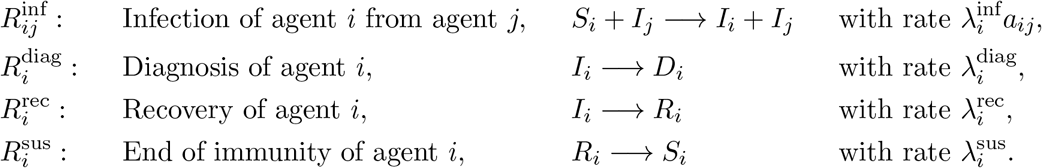

The SI model contains reaction 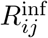, the SID model contains 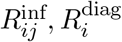, the SIDR model contains 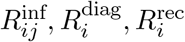 and the SIDRS model contains all listed reactions.

Transitions between compartments are primarily spontaneous; that is, they depend solely on the individual agent’s state. However, transitions from S to I are contact-driven and depend on both the agent states and the network structure.

Moreover, agents can make transient contact adjustments based on the epidemic’s progression, governed by the rate 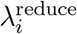. These changes depend on the epidemic’s state, as agents may be aware of the number of diagnosed individuals, causing 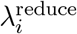 to scale with *D*(*t*). If an agent *i* adjusts its behavior, we set its contact creation rate to 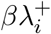. This behavior is reversed at a rate 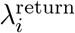, which scales inversely with the diagnosed compartment’s size.

### 2.3 Exact stochastic simulation of the system

At each time *t* the set of possible transition events is denoted by Ω(*t*). The set of transitions can be portioned into ‘network events’, ‘contact dependent events’ and ‘spontaneous transition events’. In our model the contact dependent events are infection events. The network events consist of one event per edge which is either a creation event if *a*_*ij*_(*t*) = 0 and a deletion event if *a*_*ij*_(*t*) = 1. The infection events contain the infection of the susceptible agents. All other transitions between the compartments are collected in the transition events, which are diagnosis or recovery. The waiting times for all events are exponentially distributed. The parameter is the corresponding rate. This implies that the time to the next event in the system is exponentially distributed with parameter

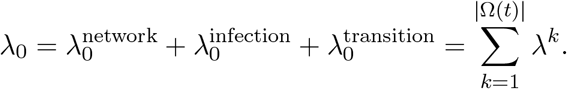

The *k*th reaction is chosen proportionally to its rate, i.e. *λ*^*k*^*/λ*_0_. Executing the transition and updating the contact network and the epidemic state accordingly gives the state of the system at a new time step. This procedure can be repeated until a time *T* is reached or a specific criterion such as *I*(*t*) + *D*(*t*) = 0 (pandemic over) is met, see Fig 1 A.

**Figure 1.**
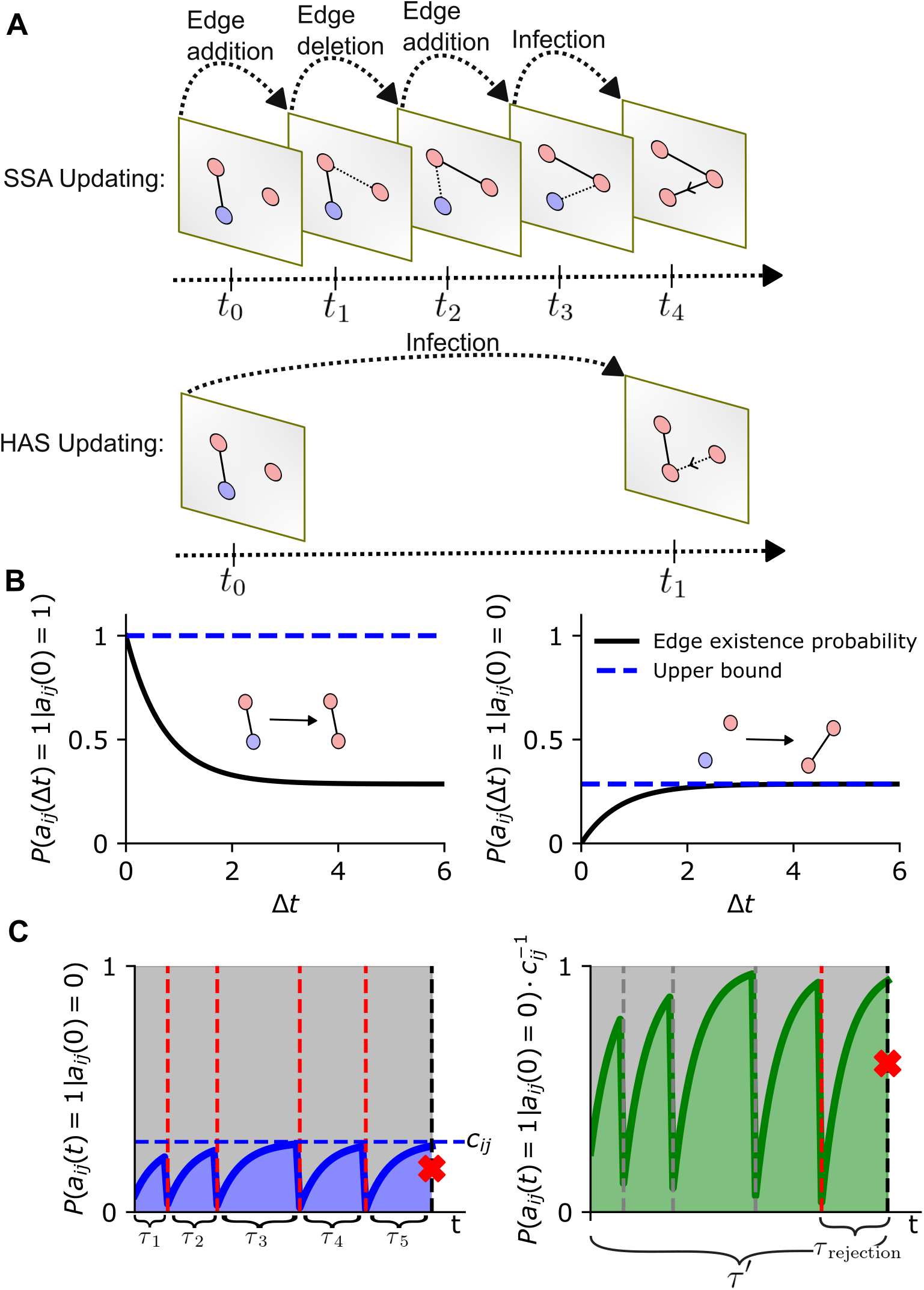
Methodological overview. **A**.Updating steps of the infection network using SSA (top) and HAS (bottom). SSA updates all network changes, whereas HAS jumps directly to the first infection event. **B**. Edge existence probability after Δ*t* when the edge exists at time *t*_0_ (left) and when it did not exist at time *t*_0_ (right). **C**. All rejection steps for edge *a*_*ij*_ until the transition is accepted. The blue area depicts the edge existence probability and the green area depicts the rescaled edge existence probability, while the dashed horizontal line shows the upper bound of the edge existence probability *c*_*ij*_. The left shows the process with *naive* upper bound *B*_0_ from (2), and the right shows the process with *tight* upper bound *B* from eq. (3). On the right, all rejection steps are bypassed, and the time to the last rejection step 13 must be sampled after moving to the infection event. Red vertical lines show rejected time points. The grey vertical lines show rejected time points that where skipped.

This algorithm is exact, i.e. all distributions and moments obtained converge to their exact values. However, the computational complexity is dominated by the contact transitions. The expected running time of SSA is given by the expected number of updating steps multiplied with the expected running time per updating step [54]. The expected number of updating steps is the inverse of the expected time steps, which again is the inverse of the expected sum of reaction propensities. The time steps are inversely proportional to the sum of the rates. The expected running time of SSA is given by

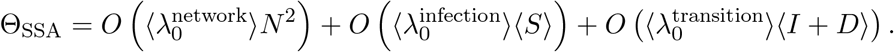

The first term corresponds to the total effort of simulating the network, the second term comes form the effort of infections and the last term accounts for transition events (I to D and {I, D} to R). The expected effort of an updating step of the network or an infection is *N*, since both actions require two agents, that need to be found. The expected size of the sum of the rates are

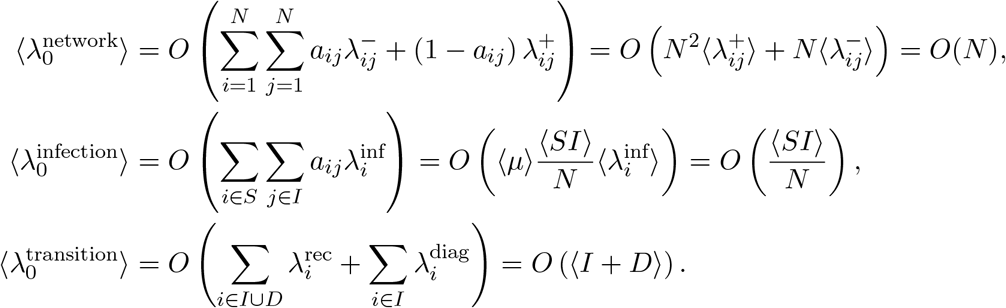

Note that the time step for the network process consistently scales as *O*(1*/N*). In contrast, the time step of the epidemic propagation process scales as *O*(1*/*(*N* − *R*), which approaches *O*(1), as the number of recovered individuals increases. This means that the expected running time of SSA scales according to the network process

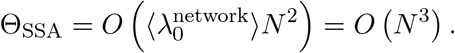

### 2.4 High acceptance sampling (HAS) for adaptive networks

In the following section, we derive an exact sampling method for modeling disease spread on adaptive networks. The running time of the method scales not with the network process, but rather with the infection process, as *O*((*N* −*R*(*t*))*N* ^2^). To achieve this, we employ acceptancerejection sampling [55], which reduces simulation effort while generating exact trajectories. In the acceptance-rejection method, an upper bound *B* on the sum of the transition rates *λ*_0_ is used to determine the next time step. The *k*th reaction is selected with rate *λ*^*k*^*/B*. Since *B* is an upper bound on the sum of all rates *λ*_0_, there is a non-zero probability that none of the transitions will be selected. In this case only the time is updated and no transition occurs. This method is particularly useful when the rates are time-dependent. We introduce a tight upper bound *B* to maximise the number of acceptance steps.

The state of the network influences the contact-dependent infection rates. Instead of realising the process governing the existence of the edge *a*_*ij*_ over the entire simulation, we sample the state on demand using the probability of edge existence *P* (*a*_*ij*_(*t*) = 1). We have the following Markovian master equation

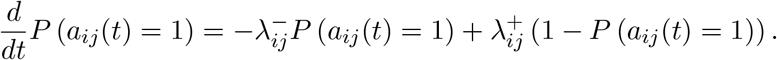

Assuming *y*_0_ ∈ {0, 1} as initial condition, this ODE has the two solutions:

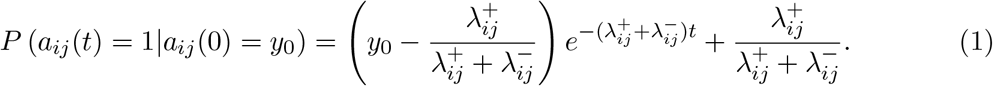

This allows on-demand sampling of the edge state at any time.

#### Naive upper bound

If we assume that an infection can spread from any infected or diagnosed agent to any susceptible agent, we obtain the following *naive* upper bound *B*_0_ on the sum of the propensities of the transitions of the spreading process (‘contact-dependent-’ and ‘spontaneous transitions’)

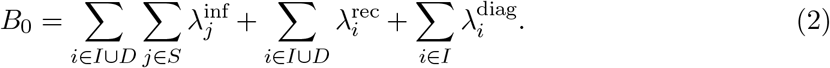

The time step *τ* is exponentially distributed with parameter *B*_0_, and transition *k* is selected proportional to its rate *λ*^*k*^*/B*_0_. If the selected transition event is an infection event from agent *i* to agent *j*, we compute the edge existence probability of *a*_*ij*_ by evaluating *P* (*a*_*ij*_(*t* + *τ*) = 1|*a*_*ij*_(**Δ**_*ij*_) = *y*_0_), where **Δ** contains the times when the edges were last observed. If *P* (*a*_*ij*_(*t* + *τ*) = 1|*a*_*ij*_(**Δ**_*ij*_) = *y*_0_) is greater than a uniformly random number, agent *j* is infected. Otherwise, the infection is rejected and we set **Δ**_*ij*_ → *t* + *τ*.

#### Tight upper bound

The probability of edge existence *P* (*a*_*ij*_(*t*) = 1 |*a*_*ij*_(0) = *y*_0_) will generally be small, since 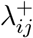 is small compared to 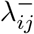, which results in a sparse contact network. To reduce the computational overhead of many rejection steps, we need to minimise the propensity bound *B*_0_. When the pair (*i, j*) is selected, the infection spreads with probability *P* (*a*_*ij*_(*t*) = 1|*a*_*ij*_(0) = *y*_0_). This means that the edge *a*_*ij*_ is equipped with two events, spreading and rejection, which have the rate 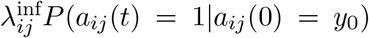 and 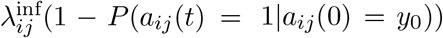, respectively. Note that the previous state of the edge **Δ**_*ij*_ is only 1 if the edge was initialised to exist at the beginning of the simulation. Therefore we assume that no edge exists at *t* = 0 and set *y*_0_ = 0. The function *P* (*a*_*ij*_(*t*) = 1 *a*_*ij*_(0) = 0) is bounded above by 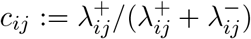, see Fig 1 C, which means that the rate of acceptance of the spread over *a*_*ij*_ is bounded by 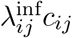. This introduces the tight constant bound for the pandemic transition rates

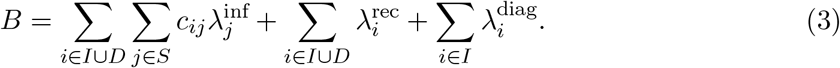

We use this upper bound to generate larger time steps and reduce the rejection probability compared to the process with the *naive* upper bound *B*_0_. With the tight upper bound, many rejection steps are leaped over. Focusing on a single edge *a*_*ij*_, we need the time between the last rejection of an infection along *a*_*ij*_ and the current time *t*. This time is exponentially distributed with parameter 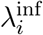, but the actual sampled period *t* − **Δ**_*ij*_ is exponentially distributed with parameter 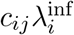. Therefore, we sample an additional time step 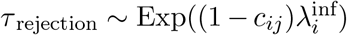 and obtain

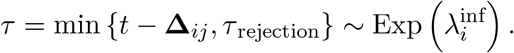

The edge existence probability is then evaluated using a time span of length *τ* (*P* (*a*_*ij*_(*τ*) = 1|*a*_*ij*_(0) = 0)). This ensures that the edge existence probability still accounts for leaped rejection steps, see Fig 1 D.

Algorithmically, a step of HAS is performed in the following way:

1. Calculate the *tight* upper bound *B* using eq. (3)
2. Sample the time increment *τ* ∼ Exp(*B*) and set *t* → *t* + *τ*
3. Sample the next event with probability *λ*^*k*^*/B*
4. If the next event is a ‘spontaneous transition’, i.e. *diagnosis* or *recovery*, execute the event and go to step 1.
5. Else edge (*i, j*) is selected for a possible ‘contact-dependent transition’, i.e. infection. Sample 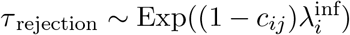 and set *τ* = min{*t* − **Δ**_*ij*_, *τ* _rejection_}. If *P* (*a*_*ij*_(*τ*) = 1|*a*_*ij*_(0) = 0) ≥ *u* for *u* ∼ *U* (0, 1), infect agent *j*, else set **Δ**_*ij*_ → *t*. Go to step 1.

The calculation of *B* requires *O*(*N* ^2^) steps, but since only a single rate changes per time step, a subsequent update of *B* can be done with constant effort *O*(1).

HAS is exact, a proof can be found in the Supplementary Materials. For each infection event, the acceptance probability for a step of size *τ* is given by

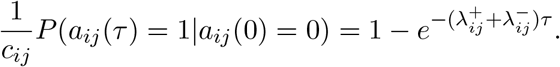

The step size is exponentially distributed with parameter 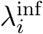 probability is given by

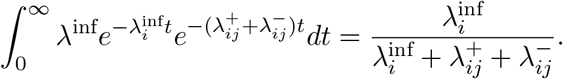

This is dominated by the infection probability (infection before edge dissolving).

## 3 Comparison of HAS and SSA

### 3.1 Numerical evaluation of exactness

We compare the outcomes of SID processes on adaptive temporal networks (model details in *Methods* section, simulated using both the SSA and the HAS methods. As can be seen in Fig 2, the first and second moments, as well as the distribution of agents across compartments at the end time, are indistinguishable between the two methods, confirming the exactness of the HAS method (see Fig 2). Sampling 100,000 trajectories took 1,768 seconds with SSA, whereas it required only 59 seconds with HAS.

**Figure 2.**
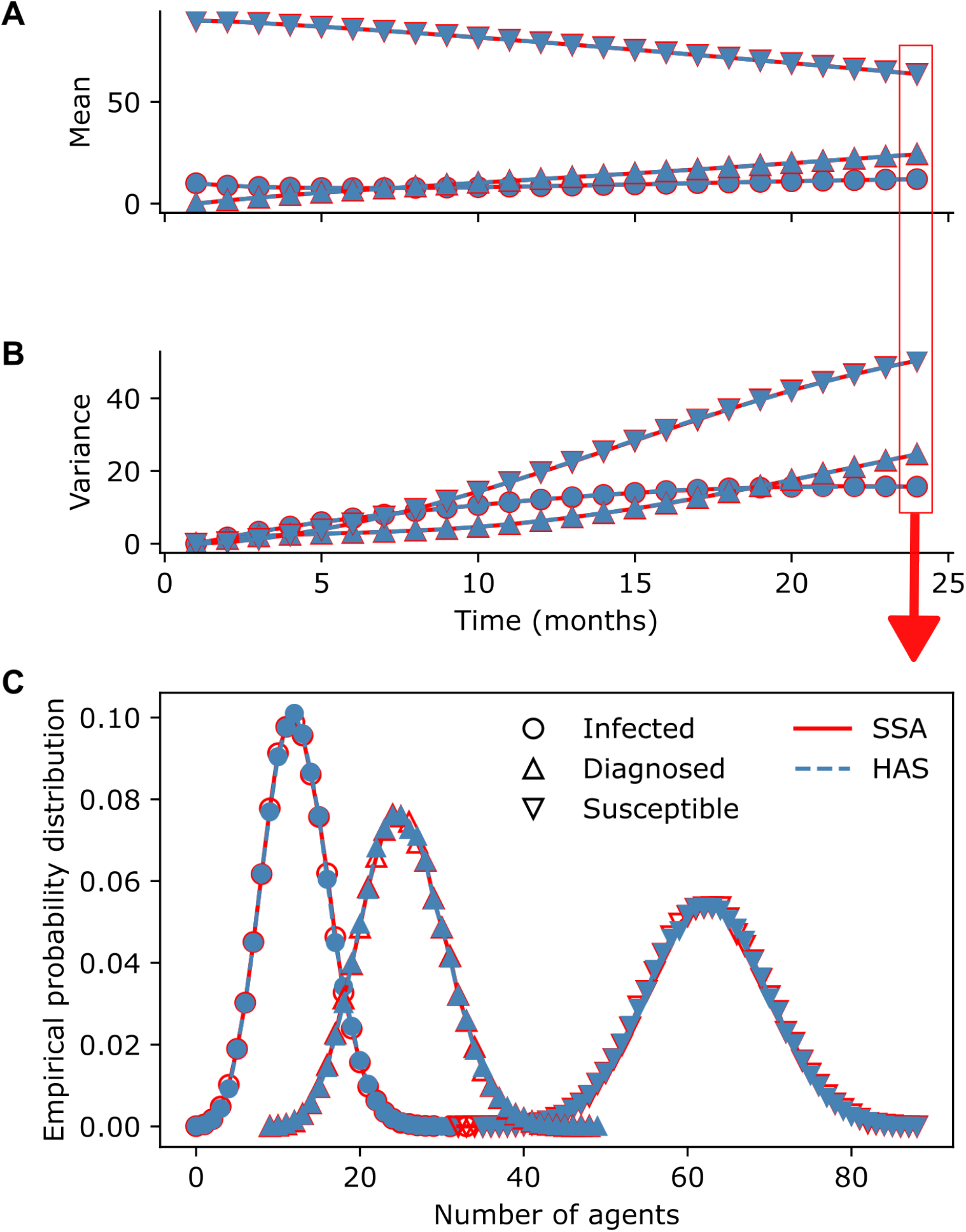
Comparison of numerical results for an SID model simulated with SSA vs. HAS. The network system consists of *N* = 100 agents with an initial infection size of *I*(0) = 10. Infection rates are distributed 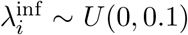, diagnosis rates 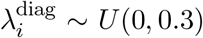. The expected number of contacts per time step is sampled from an exponential distribution with parameter 1, and each value is shifted by 1 to eliminate agents with 0 contacts. Each contact is expected to last for one time step and one time step. A total of 100,000 trajectories were sampled for each method. **A**. Mean number of agents per compartment (SID) per time step. **B**. Variance of the number of agents per compartment per time step. **C**. Distribution of agents per compartment at the end of the simulation *T* = 24.

### 3.2 Numerical comparison of sampling speed

In order to quantify the speed difference between SSA and HAS, we examined the runtimes of both methods in scenarios with varying parameters. The varied parameters are system size (Fig 3 A), edge existence probability 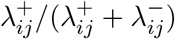 (Fig 3 B) and infection probability 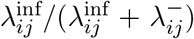 (Fig 3 C). Simulations with SSA become infeasible for larger systems or systems with many contacts. Although HAS also grows polynomially with *N*, its exponent is lower than for SSA. In the analyzed simulation setup, sampling 1,000 trajectories of a system with 100,000 agents using HAS is faster than sampling a single trajectory of the same system using SSA. As the infection probability approaches 1, the runtimes of both methods converge. In this limit, the network dynamics completely determine the spreading, since every contact lead to infection. But the runtime of HAS remains constant for a wide range of infection probabilities.

**Figure 3.**
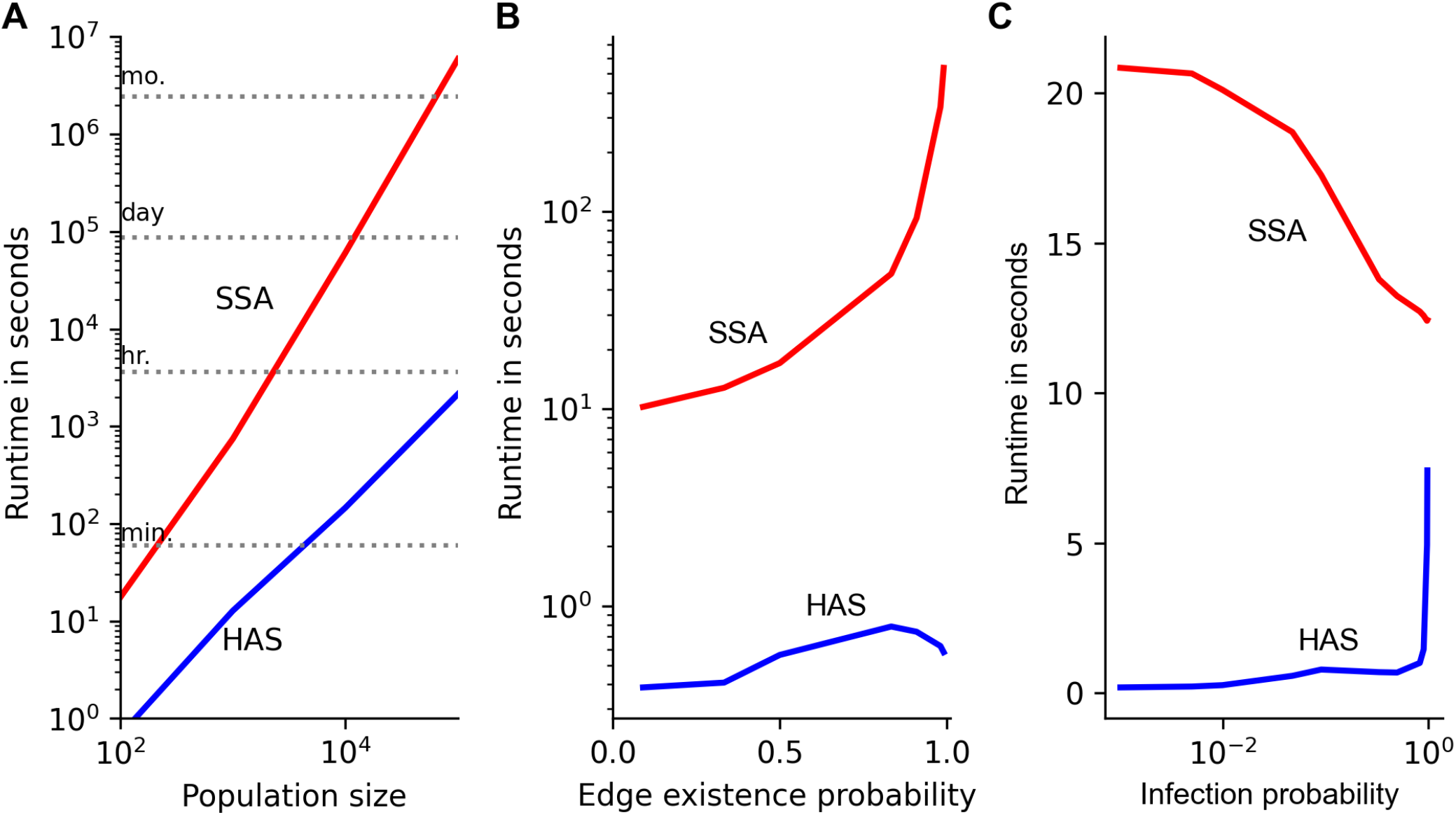
Comparison of computational speed between HAS and SSA. The base process involves *N* = 100 agents, starting with an initial infection size of *I*(0) = 10. Infection and diagnosis rates are homogeneous, set at 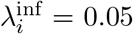 and 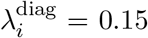, respectively. The expected number of contacts per time step is drawn from an exponential distribution with parameter 1, ensuring no agents have zero contacts by shifting each value by 1. Each contact persists on average for one time step. We sampled 1,000 trajectories for each parameter and method up to an end time of *T* = 24. **A**. System size varied from 100 to 100,000 agents. **B**. The probability of edge existence, defined as 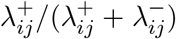, was adjusted between 0 and 1 while keeping the edge deletion rate constant and modifying *λ*^+^. **C**. The infection probability, calculated as 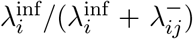, was varied by holding the edge deletion rate constant and altering the infection rate *λ*^inf^. The SSA algorithm reaches completion once every agent is infected, benefiting from a reduction in running time when the infection probability is close to 1. Computations were conducted on a Xeon Skylake 6130 with 3GB of RAM [66]

### 3.3 Theoretical sampling speed of HAS

Similar to the complexity of SSA, the expected running time of HAS is given by

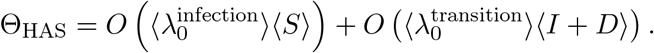

The first term is given by the infection events, which require an effort of *N* ^2^ (need to find two nodes in a list of size *N*) and the second term is given by the other spontaneous transitions that are independent of contacts. The expected size of the average transition rate is the same as with SSA, but the expected network infection rates are different

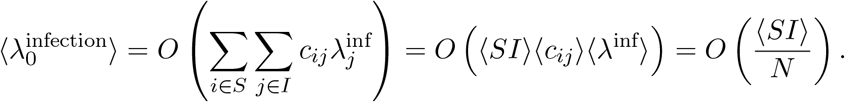

Identifying the infection event has size ⟨*SI*⟩ and hence the total effort of HAS scales with the infection process

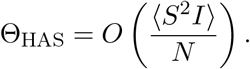

If we assume that the compartments are on average of size *O*(*N*), the expected effort for HAS becomes *O*(*N* ^2^), which is *O*(*N*) faster than SSA. However, the examples in Fig 3 suggest that the speed increase could be higher. Most of the time either *S* or *I* is of size *O*(*N*) and the other is smaller, which can move the speed increase closer to *O*(*N* ^2^).

## 4 Examples of spreading processes on adaptive networks

### 4.1 Transient behaviour changes in response to infection incidence

We study the spread of an infection in an SIRD model on three types of contact network, each with the same mean node degree but differing in degree distribution: uniform, exponential, and Poisson distributions. Each network consists of *N* = 1, 000 agents, with each agent having an average of 10 contacts per week (Fig 4A). An S-I contact has a 55% probability of transmitting the disease. Half of the infected agents are getting diagnosed, at which point the diagnosed agents lose all contacts until recovery. Recovery occurs, on average, 5 days post-infection, corresponding to a recovery rate of *λ*^rec^ = 1*/*5.

**Figure 4.**
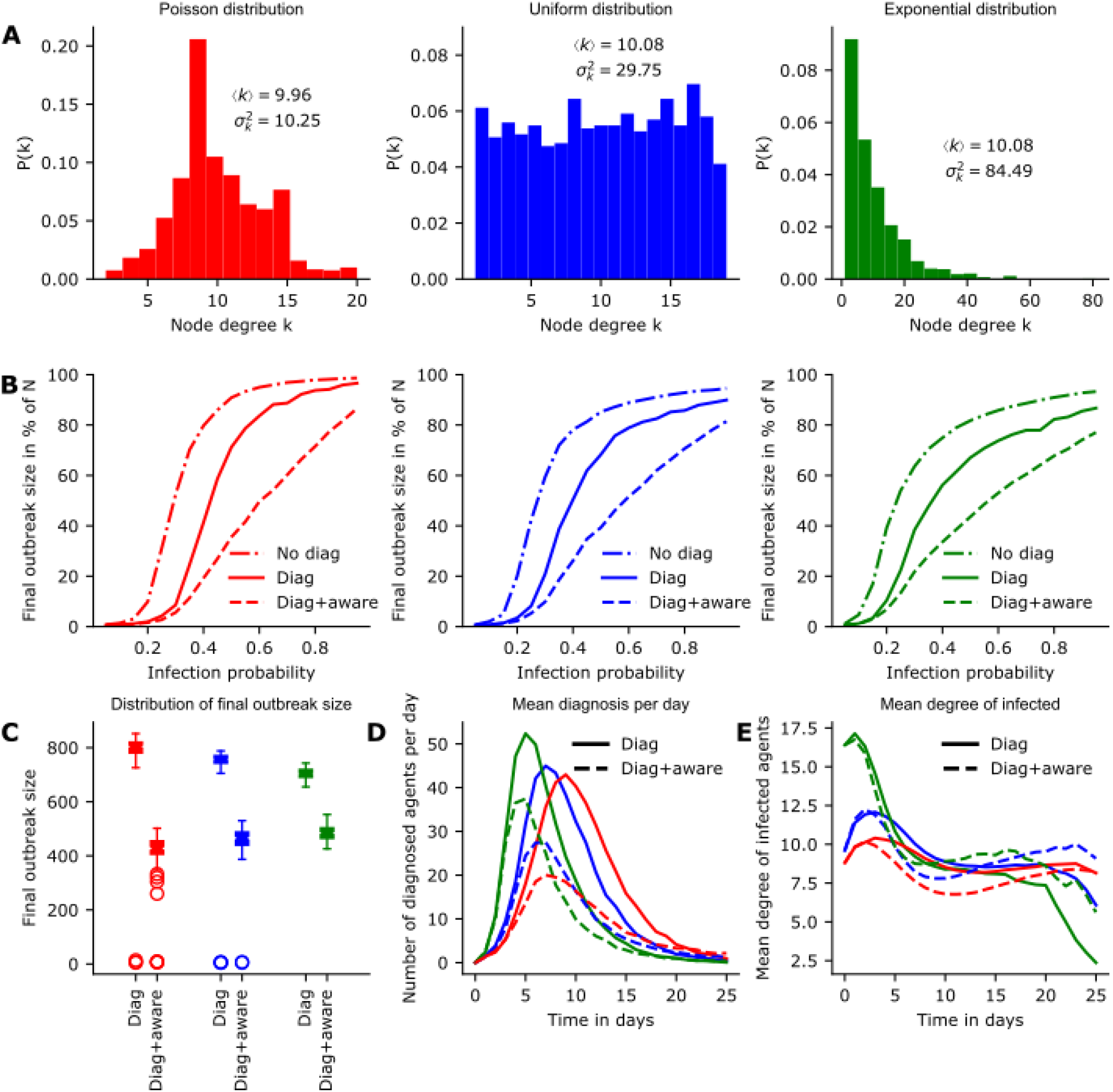
Transient behaviour changes on different network types. We simulated an outbreak on an SIDR model across different network degree distributions and scenarios. In the first scenario, agents reduce their contacts after diagnosis, in the second scenario undiagnosed agents are *aware* and additionally changed their contact behaviour in response to infection incidence (number of diagnosed agents). The populations consist of *N* = 1, 000 agents, an S-I contact has a 55% probability of transmitting the disease, the diagnosis probability is 50% and recovery occurs on average 5 days post-infection. **A**. Degree distributions for the three networks. While the mean degree ⟨*k*⟩ remains consistent across all setups, the degree variance 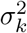 differs. **B**. Final outbreak size for different values of infection probability for the three network types and scenarios as % of *N*. The colors correspond to the colors of the distributions above. Dotted lines represent the aware population, dash-dotted lines show a scenario without diagnosis as a control. **C**. Distribution of final outbreak sizes as percent of the total population after 100 simulations for each network and scenario. **D**. Mean number of diagnoses per day for each network and population type. **E**. Timeline depicting the mean degree of infected agents for each network and population type over the simulation course.

We study two different scenarios per network: (i) a ‘naive’ population and (ii) an ‘aware’ population. In the scenario with the naive population, agents only change their contact behavior upon diagnosis. In contrast, the aware population consists of susceptible agents who can spontaneously reduce their contacts and mimic a diagnosed agent. The aware population has access to information about the number of diagnosed agents, *D*(*t*). As more agents become diagnosed, the remaining susceptible agents reduce their contact behavior more quickly (see Supplementary Fig S1). This behavior was observed, for example, during the 2022 mpox outbreak in communities of men who have sex with men [6]. We define the rates *λ*^reduce^ and *λ*^return^ to represent the reduction of contacts and the return to normal behavior, respectively:

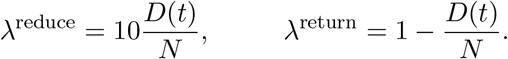

We simulate 100 trajectories per network and population type. For the naive population, the epidemic curve, defined as the number of new diagnosed cases per day, is influenced by the variance in node degrees across the network (Fig 4D): In an exponential network, the disease initially spreads rapidly as a small number of high-degree nodes become infected. Following this phase, the outbreak shifts from spreading among densely connected individuals to those with fewer connections. This transition eventually drives the epidemic below the reproductive threshold *R*_0_, which is dependent on the number of susceptible-infected (S-I) contacts (Fig 4E). In contrast, the disease spreads more slowly in the Poisson and uniform networks, as these lack many high-degree nodes or superspreaders. However, the final outbreak size is larger in these networks (Fig 4C), as the proportion of low-degree agents (or dead ends) is smaller.

For the ‘aware’ populations, the impact of spontaneous contact reduction in response to infection incidence is more pronounced in networks with slower disease spread. Again, in the exponential network, the outbreak rapidly infects most agents who can transmit the disease. Meanwhile, in the low-variance networks (uniform and Poisson), contact reduction in response to incidence is more effective, decreasing the average degree of infected agents. As a result, the final outbreak sizes become comparable across all three network types (Fig 4C).

### 4.2 Population-wide simultaneous behaviour reduction

Lastly, we use HAS to study population-wide outbreak containment strategy within an SIDRS model. The size of the system is *N* = 10, 000 with initially *I*(0) = 10 infected agents. In this example, we set the diagnosis, infection and recovery rates to be homogeneous, given as *λ*^inf^ = 0.1, *λ*^rec^ = 1*/*10 and *λ*^diag^ = 1*/*20, respectively. Rates are given per day which means that all agents are expected to test once every twenty days for infection and infected agents remain infectious for an average of ten days to mimic COVID-like dynamics [56]. Contacts are exponentially distributed with parameter 4, shifted by 1 to remove zero contact agents, yielding an average of 5 contacts per day. Recovered agents are becoming susceptible again, on average after three months [56]. The model follows SIS-type dynamics [57]: after exponential growth the epidemic eventually reaches an equilibrium and becomes endemic. In our SIDRS model, we evaluate a population-wide outbreak containment strategy that depends on the ten-day incidence rate. If this rate reaches a critical value cv_on_ all agents must reduce their contacts to the bare minimum, i.e. 1% of the initial value. This measure is lifted once the ten-days incidence rate drops below a second critical value cv_off_. This strategy was, for example, used in many European countries from 2020 onwards to regulate opening and closing of public places during the COVID pandemic. There are three limit cases based on the choices of cv_on_ and cv_off_: if cv_off_ is very low, the pathogen will eventually die out (Fig 5B). If cv_off_ is higher than the endemic equilibrium, the system may reach the endemic point (Fig 5C). If the endemic point is not contained within the interval [*cv*_off_, *cv*_on_), the outbreak state becomes periodic (Fig5D).

**Figure 5.**
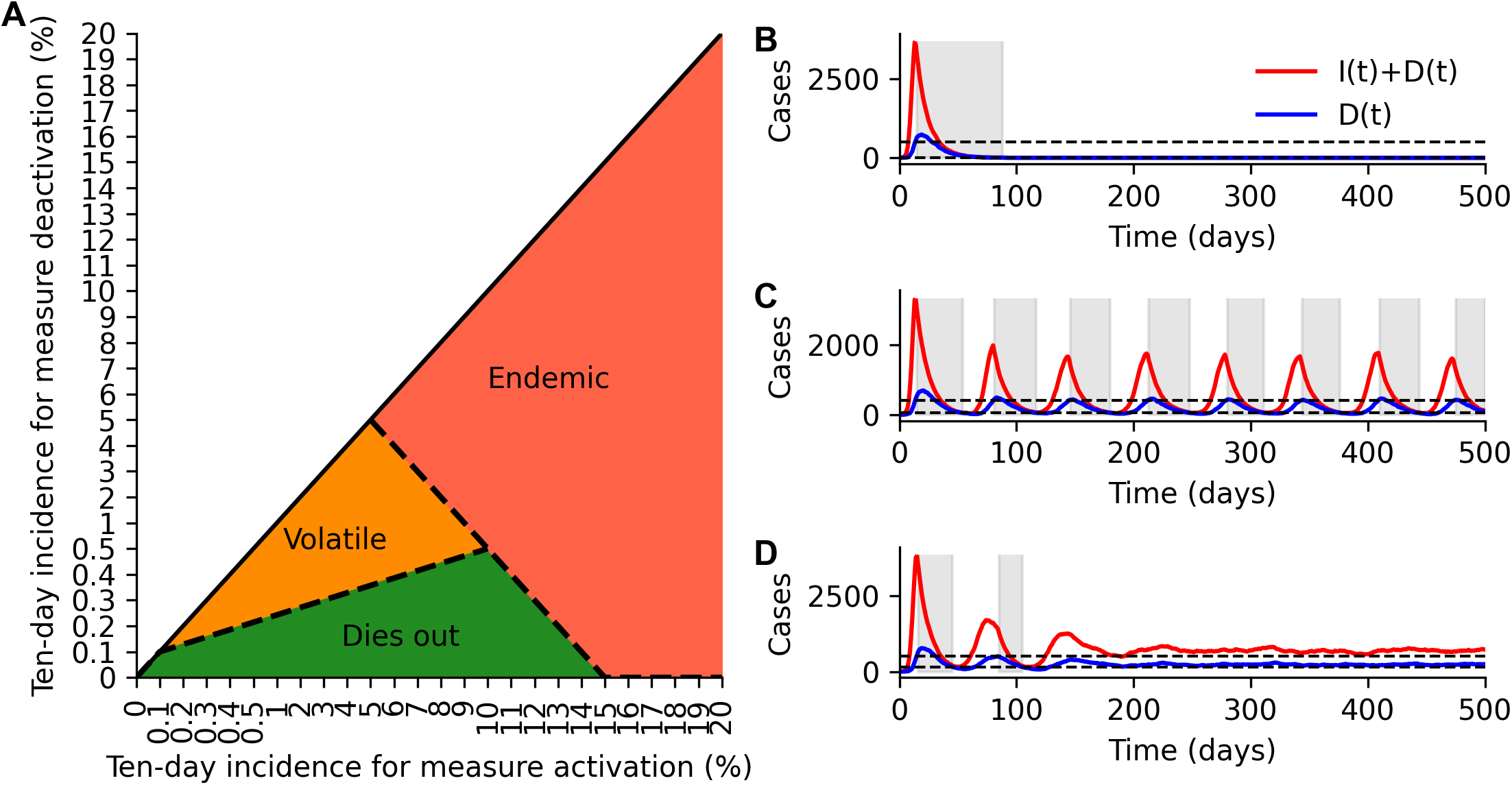
Limit cases for population-wide containment strategy. The system contains *N* = 10, 000 agents. The spreading process is SIDRS and diagnosis, infection and recovery rates are given as *λ*^inf^ = 0.1, *λ*^rec^ = 1*/*10 and *λ*^diag^ = 1*/*20, respectively. Recovered agents are becoming susceptible again on average after three months. Contacts are exponentially distributed with parameter 4, shifted by 1 to remove zero contact agents. The containment measure is activated when the ten-day incidence rate exceeds a certain threshold and deactivated when it falls below another threshold. Different combinations of these thresholds result in three distinct limit cases. **A**. Values of (*cv*_on_, *cv*_off_) and resulting limit case that predominantly occurs with the parameter set up. **B**. Representative trajectory of a pathogen extinction outcome, **C**. Representative trajectory of periodic alternation between the thresholds, or **D**. realization of endemic equilibrium. The number of diagnosed is shown in blue, and the total cases are shown in red. Shaded areas indicate when the containment measure is active.

## 5 Discussion

We have introduced the high acceptance sampling (HAS) method for simulating stochastic processes on adaptive networks. The method can be applied to simulate a number of different agent-based models, whose transitions can be separated into ‘contact events’, ‘contact-dependent transitions’ and ‘spontaneous transitions’. The core idea of the algorithm hereby is to consider the impact of the underlying contact dynamics on the ‘contact-dependent transitions’, without having to simulate the contact dynamics explicitly. Consequently, the performance of HAS is unaffected by the time scale at which contact dynamics occur, unlike SSA which becomes infeasible for rapid contact dynamics (Fig 3B). In cases with high edge existence probabilities 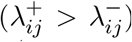 the computational efficacy of HAS even increases as the rejection probability for ‘contact-dependent transitions’ tends to zero. As the probability of ‘contact-dependent transitions’ nears 1 (the infection probability over a temporal edge in our examples), the computational effort for both SSA and HAS converge (Fig 3C). Notably, these edge cases are determined completely by the underlying contact dynamics, and hence simpler models that only consider contact dynamics would suffice.

In summary, while HAS is exact (proof in Supplementary Materials), the algorithm achieves significant speed improvements compared to other exact methods. Previously, exact (SSA) simulations of realistic systems with around 100,000 agents have been impractical. In realistic scenarios, HAS is up to 1,000 times faster than SSA (Fig 3A) and therefore enables the exact simulation of large ABMs. Such speed enhancements are crucial because sampling numerous trajectories is necessary to study the underlying stochastic process and to obtain small confidence intervals, ensuring the reliability and accuracy of the simulation results. Additionally, parameter selection often requires performing numerous simulations across a potentially high-dimensional parameter grid, which is infeasible with classic approaches such as the SSA, and unreliable with inexact methods. While inexact methods can help reduce this overhead, they often introduce errors [47]. In many applications, the core interest is on quantifying the influence of certain parameters on the infection process. If the simulation method has large errors, it becomes difficult to assess whether changes in the simulation result are due to parameter changes or numerical errors [48]. In particular for adaptive processes, inexact methods may produce incorrect statistics on the spreading process [49].

HAS is particularly suited to study the impact of behaviour change on contagion spread. While the COVID-19 pandemic and Mpox outbreaks have highlighted how behavioural change can influence the spread of pathogens across different contact networks [6, 58, 59, 60, 61, 62], traditional epidemiological modelling does typically not include socio-behavioural aspects.

In our model, the temporary prudent behavior adopted by agents following diagnosis results in fewer opportunities for them to transmit the disease compared to opportunities for them to be infected. This implies that agents at high risk of contracting the disease from others are not necessarily the ones most likely to transmit it to others. The effects on spreading dynamics in these cases was already studied on static networks, where the indegrees of agents, representing potential sources of infection, may exceed their out-degrees or the other way around [63].

Given that HAS is based on acceptance–rejection sampling, it is well-suited to handle non-constant rates. For instance, time-dependent contact rates can be implemented with no additional computational overhead (see Supplementary Material), allowing for variations such as circadian rhythms. In addition, non-constant spread rates can be implemented to account for factors such as pathogen evolution and variations in transmissibility [64] or seasonal effects. If the waiting time distributions are different from exponential, e.g. the process is non-Markovian, the acceptance-rejection sampling still holds. However, sampling the last leaped rejection step in HAS requires knowledge about the waiting time distribution. When this distribution is given in closed form, HAS can be applied to non-Markovian processes as well.

## 6 Implementation and code availability

A python implementation of HAS can be found under https://github.com/KleistLab/HAS. The main calculations are performed in C, but the method can be called via python [65].

## Data Availability

All data produced in the present study are available upon reasonable request to the authors

## 7 Acknowledgments

Funded by the Deutsche Forschungsgemeinschaft (DFG, German Research Foundation) under Germany′s Excellence Strategy – The Berlin Mathematics Research Center MATH+ (EXC-2046/1, project ID: 390685689).

The authors would like to thank the HPC Service of FUB-IT, Freie Universität Berlin, for computing time [66].

## Notes

### Competing Interest Statement

The authors have declared no competing interest.

### Author Declarations

The study only used simulated data.

