## Supplementary Materials for "Efficient and accurate simulation of infectious diseases on adaptive networks"

### Contents

**A Exactness of HAS method** **1**

**B Supplementary figures** **4**

### A Exactness of HAS method

The aim of this section is to proof that SSA and HAS generate the same distributions, i.e. the HAS method is exact. HAS has a different way of sampling infection events compared to SSA, but all other transitions between the compartments are sampled in the same way. Hence for the sake of simplicity we consider a simple two compartment SI-model: if a susceptible agent $i$ is connected to an infectious agent, the infection spreads with rate $\lambda_i^{\text{inf}}$.

**Lemma A.1.** *Let $G(t)$ be a temporal graph with $n$ nodes. An edge between vertex $i$ and $j$ is created with rate $\lambda_{ij}^+$ and removed with rate $\lambda_{ij}^-$. The state of the edge $a_{ij}(t)$ after time $t$ has the probability density function*

$$p(a_{ij}(t)=1)=\begin{cases}\frac{\lambda_{ij}^+}{\lambda_{ij}^++\lambda_{ij}^-}-\frac{\lambda_{ij}^+}{\lambda_{ij}^++\lambda_{ij}^-}e^{-(\lambda_{ij}^++\lambda_{ij}^-)t}, & \text{if } a_{ij}(0)=0,\\ \frac{\lambda_{ij}^+}{\lambda_{ij}^++\lambda_{ij}^-}+\left(1-\frac{\lambda_{ij}^+}{\lambda_{ij}^++\lambda_{ij}^-}\right)e^{-(\lambda_{ij}^++\lambda_{ij}^-)t}, & \text{if } a_{ij}(0)=1.\end{cases}$$

*Proof.* The state of the edge $a_{ij}$ can be described by a two state Markov chain with rates $\lambda_{ij}^+$ and $\lambda_{ij}^-$. The Markov chain has the following rate matrix [1]

$$\boldsymbol{Q}=\begin{pmatrix}-\lambda_{ij}^- & \lambda_{ij}^-\\ \lambda_{ij}^+ & -\lambda_{ij}^+\end{pmatrix}.$$

The rate matrix has eigenvalues 0 and $-\lambda_{ij}^- - \lambda_{ij}^+$ with eigenvectors $(1,1)$ and $(-\lambda_{ij}^-/\lambda_{ij}^+, 1)$, respectively. This yields the following diagonal form of $\boldsymbol{Q}$ which is used to calculate the probability transition matrix [1]

$$\boldsymbol{P}(t) = e^{\boldsymbol{Q}t} = \begin{pmatrix} 1 & -\frac{\lambda_{ij}^-}{\lambda_{ij}^+} \\ 1 & 1 \end{pmatrix} \begin{pmatrix} 1 & 0 \\ 0 & e^{-(\lambda_{ij}^+ + \lambda_{ij}^-)t} \end{pmatrix} \begin{pmatrix} \frac{\lambda_{ij}^+}{\lambda_{ij}^+ + \lambda_{ij}^-} & \frac{\lambda_{ij}^-}{\lambda_{ij}^+ + \lambda_{ij}^-} \\ -\frac{\lambda_{ij}^+}{\lambda_{ij}^+ + \lambda_{ij}^-} & \frac{\lambda_{ij}^+}{\lambda_{ij}^+ + \lambda_{ij}^-} \end{pmatrix}$$

$$= \begin{pmatrix} \frac{\lambda_{ij}^+}{\lambda_{ij}^+ + \lambda_{ij}^-} + \frac{\lambda_{ij}^-}{\lambda_{ij}^+ + \lambda_{ij}^-} e^{-(\lambda_{ij}^+ + \lambda_{ij}^-)t} & \frac{\lambda_{ij}^-}{\lambda_{ij}^+ + \lambda_{ij}^-} - \frac{\lambda_{ij}^-}{\lambda_{ij}^+ + \lambda_{ij}^-} e^{-(\lambda_{ij}^+ + \lambda_{ij}^-)t} \\ \frac{\lambda_{ij}^+}{\lambda_{ij}^+ + \lambda_{ij}^-} - \frac{\lambda_{ij}^+}{\lambda_{ij}^+ + \lambda_{ij}^-} e^{-(\lambda_{ij}^+ + \lambda_{ij}^-)t} & \frac{\lambda_{ij}^-}{\lambda_{ij}^+ + \lambda_{ij}^-} + \frac{\lambda_{ij}^+}{\lambda_{ij}^+ + \lambda_{ij}^-} e^{-(\lambda_{ij}^+ + \lambda_{ij}^-)t} \end{pmatrix}$$

The transition probability $a_{ij}(0) = 0 \to a_{ij}(t) = 1$ can be found in $\boldsymbol{P}_{21}(t)$ and $a_{ij}(0) = 1 \to a_{ij}(t) = 1$ can be found in $\boldsymbol{P}_{11}(t)$. □

**Remark A.2.** *If the rates are time-dependent, for example $\lambda_{ij}^+(t)$, we obtain the following first-order, linear, inhomogeneous ode*

$$\frac{d}{dt} p\left(a_{ij}(t) = 1\right) = -\lambda_{ij}^- p\left(a_{ij}(t) = 1\right) + \lambda_{ij}^+(t)\left(1 - p\left(a_{ij}(t) = 1\right)\right)$$

$$\Leftrightarrow \qquad \lambda_{ij}^+(t) = \frac{d}{dt} p\left(a_{ij}(t) = 1\right) + \left(\lambda_{ij}^- + \lambda_{ij}^+(t)\right) p\left(a_{ij}(t) = 1\right).$$

*The solution [2, 2.1] to this ODE is*

$$p\left(a_{ij}(t) = 1\right) = e^{-\int_0^t \lambda_{ij}^- + \lambda_{ij}^+(s) ds} \left( \int_0^t e^{\int_0^s \lambda_{ij}^- + \lambda_{ij}^+(v) dv} \lambda_{ij}^+(s) ds + C \right),$$

*where $C$ is a constant depending on the initial state of the edge.*

**Lemma A.3.** *The following acceptance–rejection sampling algorithm is exact:*

1. *Calculate $B_0 = \sum_{i \in I} \sum_{j \in S} \lambda_j^{inf}$*
2. *Sample the time increment $\tau \sim Exp(B_0)$ and set $t \to t + \tau$*
3. *Select infection from $i$ to $j$ with probability $\lambda_j^{inf}/B_0$*
4. *Sample the state if the edge $(i, j)$ using $p(a_{ij}(t) = 1)$. If the edge exists, infect agent $i$, else save the state of edge at time $t$.*

*Proof.* By the independence of processes it is sufficient to focus on a single S-I edge. We fix $i$ and $j$ and show that the time until the infection spreads from $i$ to $j$ in the A–R process follows the same distribution as in the SSA process. Algorthmically, the SSA performs the following steps:

1. Let $t \leftarrow 0$.
2. Generate $\tau \sim \text{Exp}(\lambda_{ij}^+)$; let $t \leftarrow t + \tau$.
3. Generate $\tau_1 \sim \text{Exp}(\lambda_{ij}^-)$ and $\tau_2 \sim \text{Exp}(\lambda_j^{\text{inf}})$; if $\tau_2 < \tau_1$, perform infection, else set $t \leftarrow t + \tau_1$ and return to 1.

This process can be split into two independent processes: one that determines the state of $a_{ij}$ and one that determines the spreading. The rate of the latter process is $\lambda_j^{\text{inf}}(t) = a_{ij}(t)\lambda_j^{\text{inf}}$ meaning the rate is non homogeneous and depending on the state of the process governing $a_{ij}$. The infection process can be uniformized by the simple observation $\lambda_j^{\text{inf}}(t) \leq \lambda_j^{\text{inf}}$. The uniformized process with constant rate $\lambda_j^{\text{inf}}$ acceptance each step with probability $\lambda_j^{\text{inf}}/\lambda_j^{\text{inf}}(t)$ for which the edge process is evaluated [3, Example 4.2]. If the edge exists, the diseases spreads and the infection process ends. Hence, for all rejection steps of the spreading process, the edge does not exist. By Lemma A.1, we use $p(a_{ij}(\tau) = 1|a_{ij}(0) = 0) = \lambda_j^{\text{inf}}(\tau)/\lambda_j^{\text{inf}}$ to generate the acceptance probability for a step of size $\tau \sim \text{Exp}(\lambda_j^{\text{inf}})$.

□

**Theorem A.4.** *HAS is exact.*

*Proof.* Lemma A.3 ensures the exactness of acceptance-rejection method combined with the ad hoc sampling of infectious edges. Again, by the independence of processes per edge, we focus on edge $a_{ij}$ connecting infected agent $i$ to susceptible agent $j$. For the rest of this proof, we assume that $a_{ij}(0) = 0$. Using the notation from the proof of Lemma A.3, we make the following observation

$$\frac{\lambda_j^{\text{inf}}(t+\tau)}{\lambda_j^{\text{inf}}} = p\left(a_{ij}(\tau) = 1|a_{ij}(0) = 0\right) < \frac{\lambda_{ij}^{+}}{\lambda_{ij}^{+} + \lambda_{ij}^{-}} = c_{ij}.$$

We want to utilize this fact in order to decrease the rejection probability in step 3 of the algorithm in the proof of Lemma A.3. We use a different time scaling and sample $\tau' \sim \text{Exp}(\lambda_j^{\text{inf}} c_{ij})$ which means $\tau' \overset{\mathcal{D}}{=} \frac{\tau}{c_{ij}}$ where $\tau \sim \text{Exp}(\lambda_j^{\text{inf}})$ is the time scale of the algorithm from Lemma A.3. We want to accept $\tau'$ with probability $\frac{\lambda_j^{\text{inf}}(\tau)}{\lambda_j^{\text{inf}} c_{ij}}$, but

$$\frac{\lambda_j^{\text{inf}}(\tau')}{\lambda_j^{\text{inf}} c_{ij}} \overset{\mathcal{D}}{=} 1 - e^{(-\lambda_{ij}^{+} + \lambda_{ij}^{-})\tau/c_{ij}} \overset{\mathcal{D}}{\neq} \frac{\lambda_j^{\text{inf}}(\tau)}{\lambda_j^{\text{inf}} c_{ij}}.$$

Hence, we sample $\Delta \sim \text{Exp}(\lambda_j^{\text{inf}}(1 - c_{ij}))$ and note $\min\{\tau', \Delta\} \sim \text{Exp}(\lambda_j^{\text{inf}})$ and obtain

$$\frac{\lambda_j^{\text{inf}}\left(\min\{\tau', \Delta\}\right)}{\lambda_j^{\text{inf}} c_{ij}} \overset{\mathcal{D}}{=} \frac{\lambda_j^{\text{inf}}(\tau)}{\lambda_j^{\text{inf}} c_{ij}}.$$

□

### B Supplementary figures

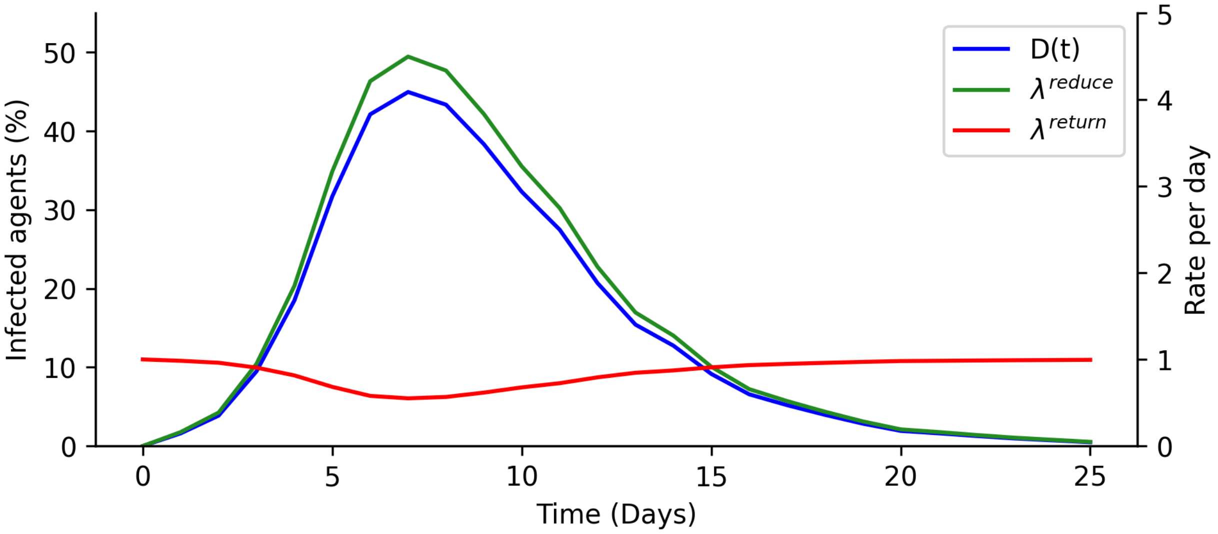


Figure S1: Illustration of contact reduction and return rates from Fig 4. The reduce rate is proportional to the number of diagnosed agents. The return rate decreases when the number of diagnosed agents peaks but is constant at other times.

[3] Søren Asmussen and Peter W Glynn. *Stochastic simulation: algorithms and analysis*, volume 57. Springer, 2007.
